# SARS-CoV-2 placental infection and inflammation leading to fetal distress and neonatal multi-organ failure in an asymptomatic woman

**DOI:** 10.1101/2020.06.08.20110437

**Authors:** Sam Schoenmakers, Pauline Snijder, Robert M. Verdijk, Thijs Kuiken, Sylvia S.M. Kamphuis, Laurens P. Koopman, Thomas B. Krasemann, Melek Rousian, Michelle Broekhuizen, Eric A.P. Steegers, Marion P.G. Koopmans, Pieter L.A. Fraaij, Irwin K.M. Reiss

**Affiliations:** Department of Obstetrics and Gynaecology; Department of Neonatology; Department of Pathology; Department of Viroscience; Department of Pediatric Infectiology, Immunology and Rheumatology; Department of Pediatric Cardiology; Department of Pharmacology and Vascular Medicine All Erasmus University Medical Center, Rotterdam

## Abstract

**Introduction:** In general SARS-CoV-2-infection during pregnancy is not considered to be an increased risk for severe maternal outcomes, but has been associated with an increased risk for fetal distress. So far, there is no direct evidence of intrauterine vertical transmission and the mechanisms leading to the adverse outcomes are not well understood

**Results:** An asymptomatic pregnant woman with preterm fetal distress during the COVID19 pandemic was included. We obtained multiple maternal, placental and neonatal swabs, which showed a median viral load in maternal blood, urine, oropharynx, fornix posterior over a period of 6 days was 5.0 log copies /mL. The maternal side of the placenta had a viral load of 4.42 log copies /mL, while the fetal side had 7.15 log copies /mL. Maternal breast milk, feces and all neonatal samples tested negative. Serology of immunoglobulins against SARS-CoV-2 was tested positive in maternal blood, but negative in umbilical cord and neonatal blood. Pathological examination of the placenta included immunohistochemical investigation against SARS-CoV-2 antigen expression in combination with SARS-CoV-2 RNA in situ hybridization and transmission electron microscopy. It showed the presence of SARS-CoV-2 particles with generalized inflammation characterized by histiocytic intervillositis with diffuse perivillous fibrin depositions with damage to the syncytiotrophoblasts.

**Discussion:** Placental infection by SARS-CoV-2 lead to fibrin depositions hampering fetal-maternal gas exchange most likely resulted in fetal distress necessitating a premature emergency caesarean section. Postpartum, the neonate showed a clinical presentation resembling a pediatric inflammatory multisystem syndrome including coronary artery ectasia, most likely associated with SARS-CoV-2 (PIMS-TS) for which admittance and care on the Neonatal Intensive Care unit (NICU) was required, despite being negative for SARS-CoV-2. This highlights the need for awareness of adverse fetal and neonatal outcomes during the current COVID-19 pandemic, especially considering that the majority of pregnant women appear asymptomatic.

## Introduction

In general SARS-CoV-2-infection during pregnancy is not considered to be an increased risk for severe maternal outcomes, but has been associated with an increased risk for fetal distress.^1^ So far, there is no direct evidence of intrauterine vertical transmission and the mechanisms leading to the adverse outcomes are not well understood. We report an intra-placental SARS-CoV-2 infection in the early third trimester diagnosed by multiple methods, including immunohistochemistry, in situ hybridization and transmission electron microscopy. Inflammation was characterized by histiocytic intervillositis with specific diffuse perivillous fibrin depositions and intervillous inflammatory infiltrates. Placental infection most likely resulted in fetal distress necessitating a premature emergency caesarean section. The neonate tested negative for SARS-CoV-2, but displayed severe multi-organ inflammatory symptoms including coronary artery ectasia for which admittance and care on the Neonatal Intensive Care unit (NICU) was required.

## Results

### Maternal

A obese primigravid woman with gestational diabetes was referred to our tertiary centre in the third trimester due to lack of fetal movements during the last 2 days. She reported general malaise, myalgia and fever a few days earlier, which resolved. At presentation to our perinatal center she had no COVID-19-related symptoms but mentioned that she come in close contact with a COVID-19-positive person. Fetal cardiotocography showed signs of severe fetal distress, including loss of beat to beat variability and repetitive decelerations, for which an emergency caesarean section was performed. Because of her medical history resembling COVID-19 related symptoms, samples for SARS-CoV-2 diagnostics (PCR and pathological analysis) were collected. (see Table 1, Figure 1). Real-time quantitative polymerase chain reaction (RT-qPCR) was performed for the detection of SARS-CoV-2 using our in-house assay^2^ or the Cobas SARS-CoV-2 test on the Cobas 6800 system © (Roche Diagnostics) depending on availability of platforms. Cycle threshold values were converted to Log10 RNA copies/mL by using calibration curves based on quantified E-gene *in vitro* transcripts as previously described.^2^ Serology was performed using the commercially available Beijing Wantai Biological Pharmacy assay. All collected samples, including placental tissue slices, tested positive for SARS-CoV-2, except for the umbilical cord blood, feces and breastmilk. Over a period of 11 days, maternal sampling was repeated (Table 1), which all remained positive for SARS-CoV-2, except for breastmilk and feces. Also, 1 day after delivery, maternal serology for SARS-CoV-2 was positive. Additional blood tests showed a slightly elevated CRP (41 mg/L) and IL-6 (11 pg/mL) levels, a positive Interferon type 1 (IFN-1) signature and normal levels of ferritin (90 ug/L), leukocytes (7.9 x10^9^/L) and D-dimers (0.40 mg/L). During admission, maternal vital parameters (temperature, heart frequency, saturation and blood pressure) remained within normal ranges and after a few days, the patient was discharged home without complaints.

**Table 1.**
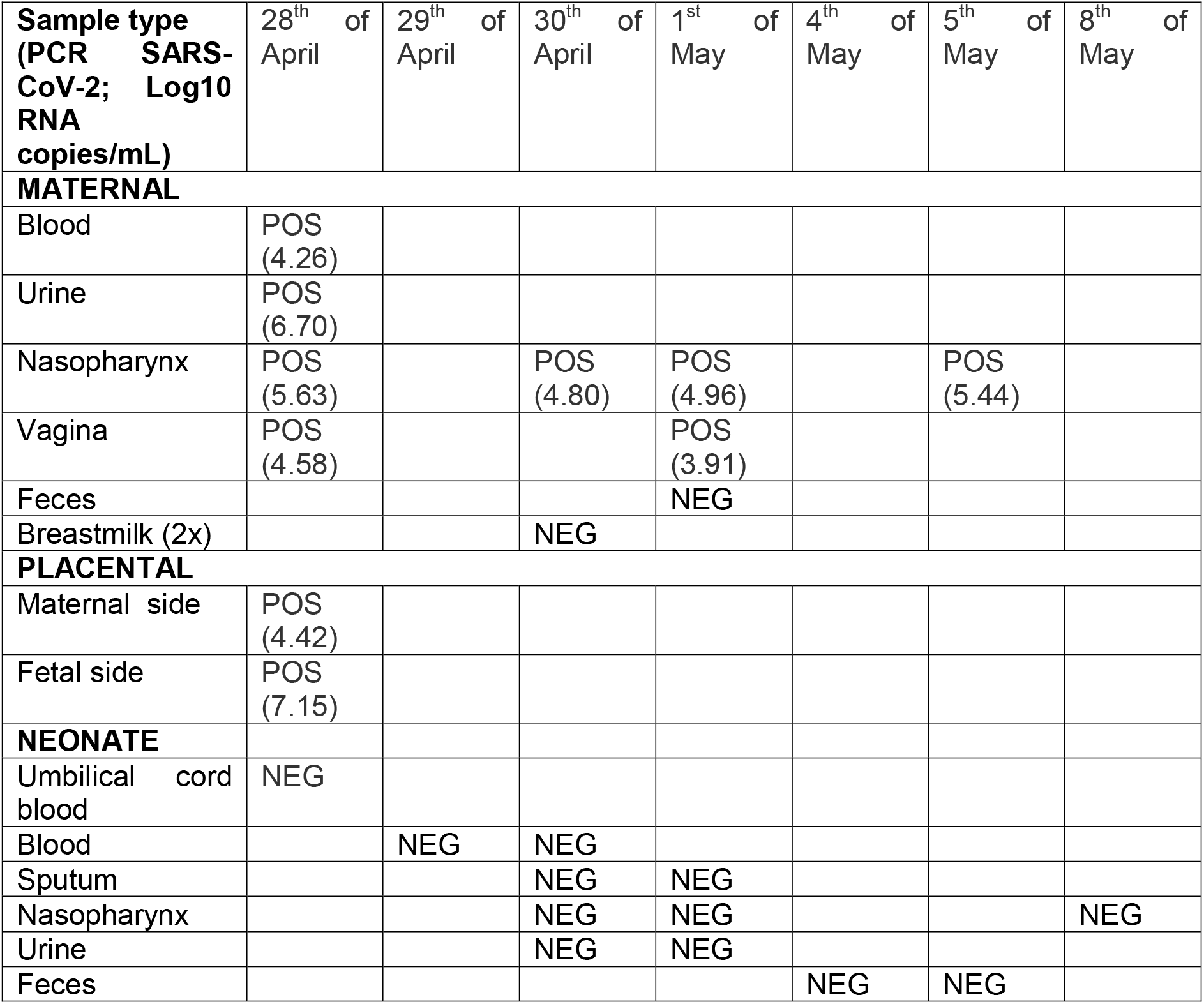
SARS-CoV-2 PCR results (log10 RNA copies/mL)

**Figure 1.**
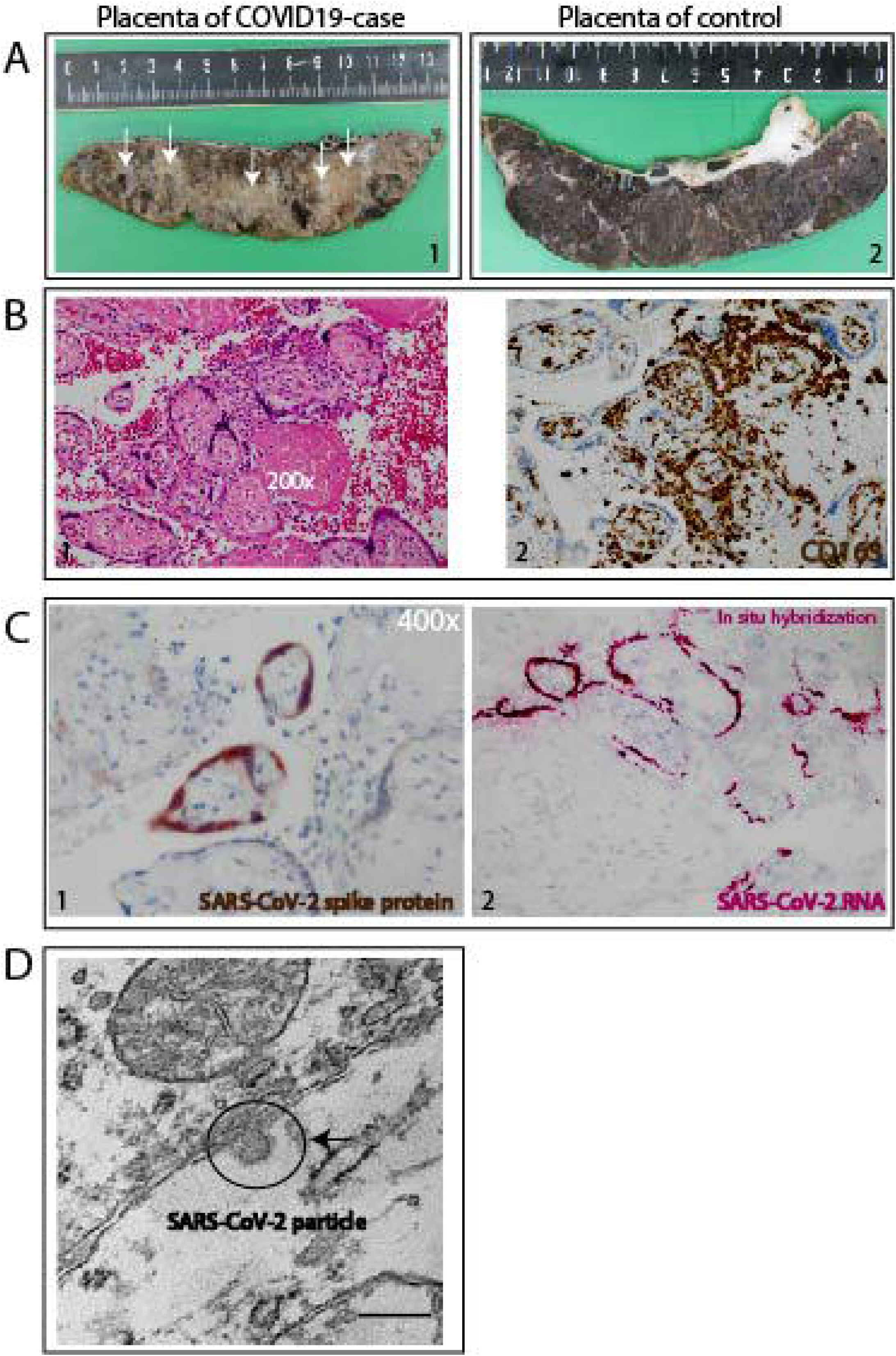
Placental syncytiotrofoblast SARS-CoV-2 infection detected by histochemical staining, specific SARS-CoV-2 RNA probe and electron microscopy. A. Gross pathology of the placenta. Case placenta slice is abnormal and shows pale trabeculae in a lattice like network (1) compared with control, age-matched placenta slice with normal appearance (2) B. Histopathology of the placenta: diffuse perivillous fibrin and an intervillous inflammatory infiltrate. 1. The intervillous inflammatory cells have a monomorphonuclear, mostly histiocytic appearance by hematoxylin and eosin staining (200x) 2. The macrophages are of the M2 phenotype as shown by CD163+-staining. C. SARS-CoV-2 infection of the syncytiotrophoblasts 1. Immunohistochemical staining SARS-CoV-2 spike protein specific antibody localizing to cytoplasm (400x) 2. In situ hybridization for SARS-CoV-2 RNA D. Electron microscopy of SARS-CoV-2 particle

### Placental examination

Gross examination showed a dense and stiff placenta with pale trabeculae in a lattice like network (arrows, Figure 1A), in line with the histological results of diffuse perivillous fibrin deposition (Figure 1B1). There was diffuse damage to the syncytiotrophoblasts associated with an intervillous inflammatory infiltrate, characterized by immunohistochemistry as M2 macrophages (CD163+ (Figure 1B2) and CD68+ (Supplemental Figure 1)), cytotoxic (CD8) and helper T-cells (CD4) as well as activated B-lymphocytes (PAX5 and CD38) (Supplemental Figure 1). No plasma-cells were detected by immunostaining for CD138 (Supplemental Figure 1). There were no signs of villous parenchyma invasion, villitis or decidual vasculopathy. Immunohistochemical investigation for SARS-CoV-2 antigen expression in combination with SARS-CoV-2 RNA in situ hybridization demonstrated predominant localization of SARS-CoV-2 in the syncytiotrophoblast cells of the placenta (Figure 1C). Electron microscopy confirmed presence of SARS-CoV-2 particles in the syncytiotrophoblast (Figure 1D), whereas villous and fetal parenchyma showed no evidence of SARS-CoV-2 infection (immunohistochemistry, in situ hybridization, or electron microscopy).

### Neonatal outcome

A female preterm infant was prematurely delivered with an Apgar score of 1, 4 and 6 at respectively 1, 5 and 10 minutes postpartum, with an umbilical cord blood pH of 6.90, a base excess of - 19 mmol/l and a birthweight of at the 75^th^ percentile. Neonatal life support was initiated on absence of spontaneous breathing and an undetectable heart rate. Because of persistent insufficient breathing and a high oxygen demand the infant was intubated, mechanically ventilated and admitted to the NICU. Chest radiography showed bilateral opacities consistent with respiratory distress syndrome for which she received repetitive dosages of pulmonary surfactant. Intra venous antibiotics were started on admission and stopped after 36 hours as blood cultures remained negative and CRP levels were low. Shortly after admission, the infant showed signs of multiple organ failure (elevated kreatinine, liver and cardiac enzymes) and developed a bilateral intraventricular hemorrhage (on the left side a grade 3 and the right side a grade 1). The patient also developed a thrombopenia and leukopenia, however differentiation showed no lymphopenia. All recovered spontaneously.

The high oxygen demand raised the suspicion of persistent pulmonary hypertension of the neonate (PPHN), which was confirmed by echocardiography. Besides the flattened interventricular septum, mild to moderate tricuspid regurgitation, a small patent ductus arteriosus with predominantly right to left shunt and a significant enlarged left main coronary artery (LMCA) was observed. To treat PPHN and systemic hypotension, inhaled nitric oxide (iNO), inotropic agents and hydrocortisone were started. Repeated echocardiograms were performed and showed an improvement of the PPHN but increasing diameter (aneurysmatic lesions) of the LMCA and left anterior descending artery (LAD). Because of the clinical presentation resembling a pediatric inflammatory multisystem syndrome, most likely associated with SARS-CoV-2 (PIMS-TS) ^3^, immunoglobulins (2 gr/kg) were administered at day 4 and aspirin was started at day 6 to prevent further coronary dilation and thromboembolic complications. ^4^ At day 14 of life only mild dilatation of the LMCA was observed.

Severely elevated levels of ferritin (14272 ug/L at day 3) as a sign of activated macrophages and significantly elevated D-dimers as sign of an endotheliitis were seen, both described in fetal inflammatory response syndrome (FIRS).^5^ An active COVID-19 infection was ruled out as all sampling of umbilical cord blood, urine, feces, blood, nasopharynx and sputum from a deep tracheal aspirate over a period of 9 days tested negative for SARS-CoV-2. Furthermore, the neonate did not develop antibodies to SARS-CoV-2. The IFN-1 signature was (repetitively) negative. In the course of the first week, inotropic support could be gradually weaned and eventually stopped at day 6 after delivery. In addition, the infant was weaned from iNO and oxygen supplementation followed by detubation at day 6 of life.

## Discussion

Our case shows that a maternal SARS-CoV-2 infection during the third trimester of pregnancy led to an adverse neonatal outcome based on a placental inflammatory reaction with subsequent dysfunction of the placenta. Remarkable is that the affected mother was in the post-infection period and had no symptoms during the event. The maternal positive IFN-1 signature indicates a strong maternal antiviral response despite absent clinical signs of SARS-CoV-2 infection during presentation. We hypothesize that the SARS-CoV-2-associated damage to the placenta will eventually lead to fetal growth restriction and distress as in our case, while fetal demise may occur when not recognized in time. Although the effects of SARS-CoV-2 infection on pregnancy and neonatal outcomes in the majority of cases seems relatively mild, complications such as miscarriage due to placental infection by SARS-CoV-2^6^, placental abruption^7^, (iatrogenic) preterm birth (21,5%), fetal distress (10.1%) and perinatal death^1^ have been reported.

We diagnosed placental inflammation caused by SARS-CoV-2 infection, based on the detection of virus infection in syncytiotrophoblasts that was colocalized with syncytiotrophoblast necrosis and a specific B-lymphocyte presentation of histiocytic intervillositis with subsequent placental failure, fetal distress and perinatal asphyxia. The observed prominent infiltrate with B-lymphocytes has not previously been described in histiocytic intervillositis ^8,9^, indicating it might be one of the histopathological hallmarks that differentiates the histiocytic intervillositis of SARS-CoV-2 infection from chronic histiocytic intervillositis of unknown origin (CHIUE).^8,9^

Currently, based on limited data, there is no evidence for intrauterine transmission of SARS-CoV-2 from infected pregnant women to their fetuses. In our case, despite the massive placental infection, all neonatal samples were negative for SARS-CoV-2, and we found no evidence for vertical transmission. This is puzzling since ACE2 receptors seem essential in transmission and infection by SARS-CoV-2 and are highly expressed on the placental maternal-fetal interface cells.^10^ We cannot rule out that the detected SARS-CoV-2 RNA by PCR on the fetal side of the placenta is caused by contamination at the time of sampling, although the number of RNA copies was substantially higher than in maternal blood. During pregnancy, the placenta forms a natural barrier against maternal viral infections although the local immune-tolerant environment might permit viral replication. The specific mechanisms allowing some viruses, such as rubella virus and the Zika virus, to cause transplacental fetal infection are not well understood.^11,12^ Placental examination of pregnant women infected with the related SARS-CoV-virus of 2002-2003 revealed increased subchorionic and intervillous fibrin with extensive fetal thrombosis.^13^ As in our case, intervillitis, a histologic characteristic of maternal hematogenous infections that can lead to congenital infection, was not observed in those placentas.^13^ Unraveling the mechanism by which the placenta prevents passage of coronaviruses onto the fetus would be of great general interest for better understanding the placental barrier function.

Placental SARS-CoV-2 infection lead to massive local inflammation with the formation of fibrin depositions, hereby decreasing the available maternal-fetal interface necessary for effective gas exchange (Figure 2) which is essential for fetal growth and development. We speculate that fetal and subsequent neonatal distress due to placental dysfunction caused by inflammation explains the clinical course. However, some clinical hallmarks (such as a dilated coronary arteries, extremely high ferritin and high D-dimer levels) were not consistent with asphyxia alone and suggest a hyperinflammatory response as seen in macrophage activation syndrome.^4,14-16^ In perinatal asphyxia, increased coronary blood flow (myocardial sparing) has been described, but enlargement of coronary arteries has not.^17^ The observed dilation of the LMCA and LAD, which is extremely rare in neonates^18^, might be a sign of endotheliitis as recently described in older children with COVID-19-related disease.^3^ In these older pediatric patients, high levels of ferritine and D-dimers were described, as seen in our neonate.^3^ Also, by use of pattern recognition receptors^5^, the fetal immune system can detect signals produced in the context of structural damage, which can activate a severe immunological response leading to FIRS. This is normally associated with severe bacterial chorioamnionitis. ^5^ In our case, it might be that placental infection by SARS-CoV-2 resulted in FIRS.

**Figure 2.**
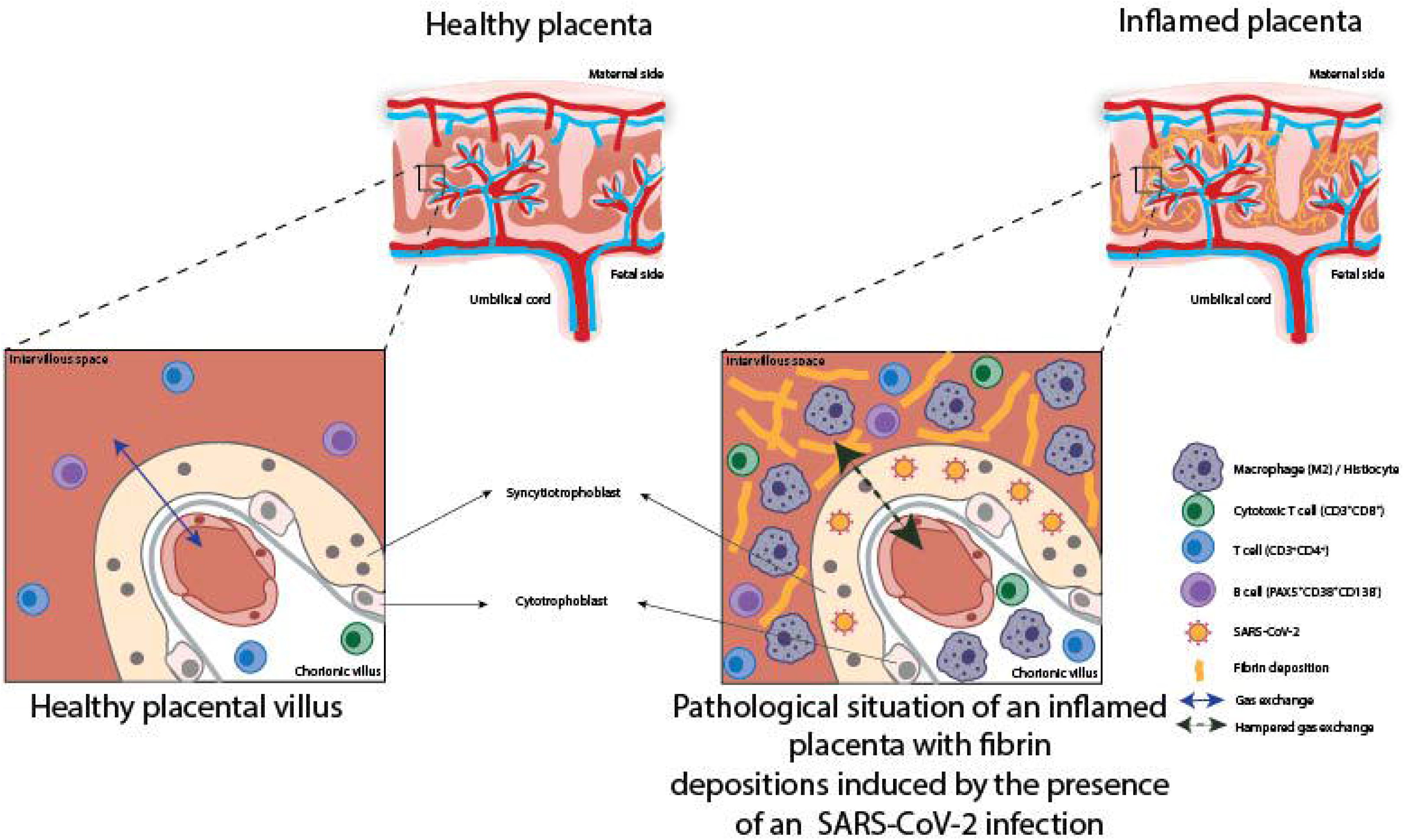
Graphical representation of a healthy placenta compared with perivillous fibrosing and placental inflammation caused by infection of SARS-CoV-2.

In conclusion we here report a SARS-CoV-2-associated inflammation of the placenta in a mother who was asymptomatic at presentation with severe fetal and neonatal consequences including coronary artery dilation. This highlights the need for awareness of adverse fetal and neonatal outcomes during the current COVID-19 pandemic, especially considering that the majority of pregnant women appear asymptomatic ^19^.

**Supplemental Figure 1: Inflammation characterization of a SARS-CoV-2-infected placenta by histochemical staining.**

A. The macrophages are of the M2 phenotype: CD68+.

B. CD3 positive T-cells are composed of mixed CD4+ T-helper & CD8+cytotoxic T-cells.

C. SARS-CoV-2-associated histiocytic intervillositis specific Pax5 positive B-cell component with specific CD38+ cells & no presence of plasma cells since CD138 only stains the viable trophoblasts and not B-cells.

## Data Availability

n.a.

